# Comparative genomics and antimicrobial resistance profiling of *Elizabethkingia* isolates reveals nosocomial transmission and *in vitro* susceptibility to fluoroquinolones, tetracyclines and trimethoprim-sulfamethoxazole

**DOI:** 10.1101/2020.03.12.20032722

**Authors:** Delaney Burnard, Letitia Gore, Andrew Henderson, Ama Ranasinghe, Haakon Bergh, Kyra Cottrell, Derek S. Sarovich, Erin P. Price, David L. Paterson, Patrick N. A. Harris

## Abstract

The *Elizabethkingia* genus has gained global attention in recent years as a nosocomial pathogen. *Elizabethkingia* spp. are intrinsically multidrug resistant, primarily infect immunocompromised individuals, and are associated with high mortality (∼20-40%). Although *Elizabethkingia* infections appear sporadically worldwide, gaps remain in our understanding of transmission, global strain relatedness and patterns of antimicrobial resistance. To address these knowledge gaps, 22 clinical isolates collected in Queensland, Australia, over a 16-year period along with six hospital environmental isolates were examined using MALDI-TOF MS (VITEK^®^ MS) and whole-genome sequencing to compare with a global strain dataset. Phylogenomic reconstruction against all publicly available genomes (*n*=100) robustly identified 22 *E. anophelis*, three *E. miricola*, two *E. meningoseptica* and one *E. bruuniana* from our isolates, most with previously undescribed diversity. Global relationships show Australian *E. anophelis* isolates are genetically related to those from the USA, England and Asia, suggesting shared ancestry. Genomic examination of clinical and environmental strains identified evidence of nosocomial transmission in patients admitted several months apart, indicating probable infection from a hospital reservoir. Furthermore, broth microdilution of the 22 clinical *Elizabethkingia* spp. isolates against 39 antimicrobials revealed almost ubiquitous resistance to aminoglycosides, carbapenems, cephalosporins and penicillins, but susceptibility to minocycline, levofloxacin and trimethoprim/sulfamethoxazole. Our study demonstrates important new insights into the genetic diversity, environmental persistence and transmission of Australian *Elizabethkingia* species. Furthermore, we show that Australian isolates are highly likely to be susceptible to minocycline, levofloxacin and trimethoprim/sulfamethoxazole, suggesting that these antimicrobials may provide effective therapy for *Elizabethkingia* infections.

**Importance:** *Elizabethkingia* are a genus of environmental Gram-negative, multidrug resistant, opportunistic pathogens. Although an uncommon cause of nosocomial and community-acquired infections, *Elizabethkingia* spp. are known to infect those with underlying co-morbidities and/or immunosuppression, with high mortality rates of ∼20-40%. *Elizabethkingia* have a presence in Australian hospitals and patients; however, their origin, epidemiology, and antibiotic resistance profile of these strains is poorly understood. Here, we performed phylogenomic analyses of clinical and hospital environmental Australian *Elizabethkingia* spp., to understand transmission and global relationships. Next, we performed extensive minimum inhibitory concentration testing to determine antimicrobial susceptibility profiles. Our findings identified a highly diverse *Elizabethkingia* population in Australia, with many being genetically related to international strains. A potential transmission source was identified within the hospital environment where two transplant patients were infected and three *E. anophelis* strains formed a clonal cluster within the phylogeny. Furthermore, near ubiquitous susceptibility to tetracyclines, fluoroquinolones and trimethoprim/sulfamethoxazole was observed in clinical isolates. We provide new insights into the origins, transmission and epidemiology of *Elizabethkingia* spp., in addition to understanding their intrinsic resistance profiles and potential effective treatment options, which has implications to managing infections and detecting outbreaks globally.

## Introduction

The genus *Elizabethkingia* (formerly *Chryseobacterium*), comprise a group of environmental bacteria that have traditionally been isolated from soil and water environments^1–4^. As opportunistic pathogens, *Elizabethkingia* spp. can cause sporadic nosocomial outbreaks and infections in immunocompromised or at-risk individuals^1,2,5–8^. Infections have been documented worldwide such as those in the Central African Republic^9^, Mauritius^10^, Singapore^11^, Taiwan^12^ and the USA^6^, suggesting a comprehensive global distribution that is yet to be fully described. Often, the source of *Elizabethkingia* spp. infection remains unclear and routes of transmission are still to be defined^2,6,9,12–16^. However, previous investigations have suggested that shared water reservoirs within hospitals may be an overlooked source of infection^1,2,17^.

As an understudied pathogen, taxonomic assignment within the *Elizabethkingia* genus is ongoing. Recently, a formal taxonomic revision using whole-genome sequencing (WGS) left the previously described species *E. meningoseptica* and *E. miricola* unchanged, while the proposed species *E. endophytica*^18^ is now considered a clone within *E. anophelis*^19–21^. Several new species, *E. bruuniana, E. ursingii*, and *E. occulta* have recently been described^3–5^. It is also now recognised that *E. anophelis*, not *E. meningoseptica*, is the primary species causing human infection, although clinical presentations may be very similar^4,13,22–24^. The remaining members of the genus are thought to be much less prevalent in human disease; however, difficulties in accurately identifying *E. miricola, E. bruuniana, E. ursingii*, and *E. occulta* from clinical specimens has hindered appropriate recognition and characterisation of these species^4^.

Common clinical presentations of *E. anophelis* infections include primary bacteraemia, pneumonia, sepsis and meningitis in neonates^7,14,22,23^. Risk factors associated with *E. anophelis* infection consist of being male, having underlying chronic medical conditions such as malignancy or diabetes mellitus, and admission to critical care or neonatal units^13,22,23,25^. Currently, approximately 80% of *E. anophelis* infections are considered hospital-acquired with mortality rates ranging from 23-26% ^22,23,25^. Similarly, *E. meningoseptica* infections also present as neonatal meningitis and/or sepsis but can also cause infections in most organ systems. Primary bacteraemia is the most common presentation, occurring more often in hospitalised patients and those with underlying co-morbidities^8,12^. The mortality rate of *E. meningoseptica* infection is between 23-41%, with higher rates in individuals where premature birth, shock or admission to a critical care unit has taken place^12,26^. To date, the largest outbreak was caused by community-acquired *E. anophelis* in Wisconsin, USA, from 2015-2016. A total of 66 individuals were infected and the outbreak spread to the neighbouring states of Illinois and Michigan^6^. Comparative genomics characterised unique mutations in an integrative conjugative element (ICE) insertion in the *MutY* gene in all infecting strains as well as a mutation in the *MutS* gene in hypermutator strains, which may have accelerated the transmission of the outbreak clone^6^.

Poorly understood intrinsic multidrug resistance (MDR) in *E. anophelis* and *E. meningoseptica* infections has led to inappropriate empiric antibiotic therapy, especially in patients with underlying co-morbidities, in critical care^12,26^ or neonatal units^1,2,8,13,15,16,27^, resulting in high mortality rates. There are currently no established minimum inhibitory concentration (MIC) breakpoints for *Elizabethkingia* spp., causing reported susceptibility rates to vary among studies. Despite interpretation differences, *Elizabethkingia* are generally considered resistant to carbapenems, cephalosporins, aminoglycosides, and most β-lactams even in combination with β-lactamase inhibitors (except for piperacillin/tazobactam). Minocycline, levofloxacin, trimethoprim/sulfamethoxazole and piperacillin/tazobactam are the most common antimicrobials that have been tested and generally demonstrate widespread susceptibility^4,6,23–25,28^. Interestingly, *in vitro* susceptibility to the Gram-positive glycopeptide vancomycin has been documented in *Elizabethkingia* spp., resulting in vancomycin being suggested as a therapy^4,29–31^. Based on these results the empiric antibacterial therapy of choice for *Elizabethkingia* spp. infections is not clear, but should ideally be guided by further MIC profiling^7,23,25,31^.

Here, we present one of the largest comparative genomic analyses of the *Elizabethkingia* genus to date, which includes 22 newly described clinical isolates and six hospital environmental isolates from Australia, a previously underrepresented geographic area. The speciation accuracy of the VITEK^®^ MS v3.2 database was assessed, in addition to a comprehensive examination of clinical isolates using both genomic data and MIC testing across 39 antimicrobials. Our results provide valuable insights into global *Elizabethkingia* relationships, speciation accuracy, transmission, the extent of intrinsic antimicrobial resistance and options for potential effective antimicrobial therapy to combat these opportunistic pathogens.

## Methods

### Ethics statement

This project was reviewed by the chairperson of a National Health and Medical Research Council (NHMRC) and registered with The Royal Brisbane and Women’s Hospital Human Research Ethics Committee (HREC) (EC00172) and was deemed compliant with the NHMRC guidance “Ethical considerations in Quality Assurance and Evaluation Activities” 2014 and exempt from HREC review.

### Isolates and initial identification

Twenty-two clinical *Elizabethkingia* spp. isolates collected in Queensland, Australia over a 16-year period (2002-2018) were included in this study (Table 1). Isolates were collected by two methods. First, laboratory database storage records from multiple public and private laboratories in Queensland were searched for *Elizabethkingia* spp. or *Chryseobacterium meningoseptica*. Second, isolates identified by current laboratory identification systems as *Elizabethkingia* spp. were collected prospectively from both private and public pathology laboratories throughout the state of Queensland between January 2017 and October 2018. All isolates were stored at -80°C with low temperature bead storage systems. Single colonies were double passaged from clinical specimens on 5% horse blood agar (Edwards Group MicroMedia, Narellan, NSW, Australia) then subjected to identification via VITEK^®^ MS Knowledge Base v3.2 (bioMérieux, Murarrie, QLD, Australia) which is inclusive of *E. anophelis, E. miricola* and *E. meningoseptica*.

**Table 1.** *Elizabethkingia* spp. isolates and associated speciation information included in the current study. Strain EkQ11, highlighted in purple represents a species identification error according to the VITEK^®^ MS Knowledge Base v3.2.

| Isolate ID | Age (y) | Date collected | Collection site | VITEK MS v3.2 ID | Whole Genome ID |
| --- | --- | --- | --- | --- | --- |
| EkQ1 | 1 | 2017 | Sputum | *E. miricola* | *E. miricola* |
| EkQ3 | 43 | 2017 | Sputum | *E. anophelis* | *E. anophelis* |
| EkQ4 | 78 | 2017 | Blood | *E. meningoseptica* | *E. meningoseptica* |
| EkQ5 | 59 | 2017 | Blood | *E. anophelis* | *E. anophelis* |
| EkQ6 | 17 | 2018 | Bronchoalveolar lavage | *E. anophelis* | *E. anophelis* |
| EkQ7 | 69 | 2018 | Blood | *E. anophelis* | *E. anophelis* |
| EkQ8 | 0 | 2018 | Urine | *E. anophelis* | *E. anophelis* |
| EkQ10 | 34 | 2018 | Sputum | *E. miricola* | *E. miricola* |
| EkQ11 | 85 | 2018 | Blood | *E. miricola* | *E. bruuniana* |
| EkQ12 | 53 | 2018 | Blood | *E. meningoseptica* | *E. meningoseptica* |
| EkQ13 | 1 | 2011 | Sputum | *E. miricola* | *E. miricola* |
| EkQ15 | 16 | 2002 | Bronchoalveolar lavage | *E. anophelis* | *E. anophelis* |
| EkQ16 | 82 | 2017 | Blood | *E. anophelis* | *E. anophelis* |
| EkQ17 | 66 | 2018 | Blood | *E. anophelis* | *E. anophelis* |
| EkM1 | Unknown | 2018 | Unknown | *E. anophelis* | *E. anophelis* |
| EkM2 | Unknown | 2018 | Unknown | *E. anophelis* | *E. anophelis* |
| EkM3 | Unknown | 2014 | Unknown | *E. anophelis* | *E. anophelis* |
| EkS1 | 80 | 2013 | Blood | *E. anophelis* | *E. anophelis* |
| EkS2 | 82 | 2015 | Blood | *E. anophelis* | *E. anophelis* |
| EkS3 | 74 | 2016 | Blood | *E. anophelis* | *E. anophelis* |
| EkS4 | 73 | 2012 | Blood | *E. anophelis* | *E. anophelis* |
| EkS5 | 66 | 2018 | Dialysis fluid | *E. anophelis* | *E. anophelis* |
| EK1 | N/A | 2019 | Toilet sink drain, Oncology Ward | *E. anophelis* | *E. anophelis* |
| EK2 | N/A | 2019 | Corridor sink drain, Infectious Disease Ward | *E. anophelis* | *E. anophelis* |
| EK3 | N/A | 2019 | Hand washing drain, Oncology Ward | *E. anophelis* | *E. anophelis* |
| EK4 | N/A | 2019 | Hand wash dink, Transplant Ward | *E. anophelis* | *E. anophelis* |
| EK5 | N/A | 2019 | Toilet handrail, Transplant Ward | *E. anophelis* | *E. anophelis* |
| EK6 | N/A | 2019 | Toilet sink, Transplant Ward | *E. anophelis* | *E. anophelis* |

Furthermore, six environmental isolates were collected in 2019 from a participating hospital via swabbing various surfaces throughout the environment (Table 1). Specimens were plated onto 5% horse blood agar and *Elizabethkingia* spp. colonies were double passaged to ensure purity then subjected to identification via VITEK^®^ MS Knowledge Base v3.2.

### DNA extraction, whole-genome sequencing and genome assembly

DNA was extracted using the DNeasy Ultra Clean Microbial extraction kit (Qiagen, Chadstone, VIC, Australia) according to the manufacturer’s instructions. Purified DNA was quantified using both the NanoDrop 3300 spectrophotometer and the Qubit^™^ 4 fluorometer (Thermo Fisher Scientific). Sequencing libraries were generated using the Nextera Flex DNA library preparation kit and sequenced on the MiniSeq^™^ System (Illumina Inc.^®^, San Diego, CA, USA) on a high output 300 cycle cartridge according to the manufacturer’s instructions. Comparative genomic analyses were performed across a large *Elizabethkingia* data set (*n*=128; Table S1), including the 28 Australian genomes generated in the current study (Table 1), to assign species and to assess intraspecific and geographical relationships among strains. Publicly available *Elizabethkingia* Illumina reads (*n*=119) were downloaded from the NCBI Sequence Read Archive database (January 2019), and *Elizabethkingia* spp. assemblies were downloaded from the GenBank database (*n*=109). Publicly available Illumina reads were quality-filtered with Trimmomatic v0.38^32^ and subject to quality control assessments with FastQC^33^, followed by downsizing using Seqtk to 40x coverage^34^. For assemblies without accompanying Illumina data, synthetic paired-end reads were generated with ART MountRainier-2016.06.05^35^. Genomes were limited to one representative per strain, and only high-quality sequence reads according to FastQC were included to avoid errors in phylogenomic reconstruction (*n*=100; Table S1). The genomes were assembled using SPAdes v3.13.0^36^ and annotated with Prokka v1.13^37^ (Table S2).

### Phylogenomic reconstruction

The comparative genomics pipeline SPANDx v3.2^38^ was used under default settings to identify orthologous, biallelic, core-genome single-nucleotide polymorphism (SNP) and short insertion-deletion (indel) characters among the 128 *Elizabethkingia* genomes. *E. anophelis* NUHP1, *E. miricola* CSID3000517120, *E. meningoseptica* G4120 and *E. bruuniana* G0146 (GenBank accession numbers NZ_CP007547.1, NZ_MAGX00000000.1, NZ_CP016378.1 and NZ_CP014337.1 respectively) were used as reference genomes for SPANDx read mapping alignment. Outputs from SPANDx were used to generate maximum parsimony trees using PAUP version 4.0a^39^ and visualised in FigTree v4.0 (http://tree.bio.ed.ac.uk/software/figtree). From the 128 genomes 127,236 SNPs and were used to construct the *Elizabethkingia* genus phylogeny (Figure 1.). Within-species phylogenies were also constructed using 121,827 SNPs from 71 genomes for *E. anophelis* (Figure 2), 135,087 SNPs from 18 genomes for *E. miricola* (Figure 3), 61,500 SNPs from 22 genomes for *E. meningoseptica* (Figure 4) and 82,680 SNPs from 10 genomes for *E. bruuniana* (Figure 5) phylogenies respectively. All phylogenies were statistically tested with 1000 bootstrap replicates. Branch support of less than 0.8 is shown in figures. To assess SNP and indel differences amongst closely related strains, the earliest collected strain was used as the reference in SPANDx, SNP and indel variants that had passed quality filtering were visualised in Tablet 1.19.09.03^40^ and Geneious Prime 2019 2.1^41^ (Table 2).

**Table 2.** Single nucleotide polymorphism and deletion differences between strains of the clonal cluster of clinical and environmental *Elizabethkingia anophelis* isolates. Clinical isolates EkQ5 (earliest collected and reference strain) and EkQ17 were collected from two different transplant patients, while Ek6 was collected from a shared handwashing sink on the transplant ward. Grey shading shows no differences, green shows similarities between EkQ17 and EK6, while blue shading highlights unique changes. The proteins affected by each mutation and the resulting amino acid changes are also shown.

| Mutation | **EkQ5**<br>2018 | EkQ17<br>2018 | EK6<br>2019 | Protein affected | Effect |
| --- | --- | --- | --- | --- | --- |
| SNP | **G** | A | A | Hypothetical Protein | A10T |
| SNP | **G** | A | A | Efflux pump membrane transporter (bepE) | S416R |
| SNP | **C** | T | T | 3-oxoacyl-[acyl-carrier-protein] synthase 2 (fabF) | R303H |
| SNP | **T** | C | C | Penicillin binding protein E (pbpE_7) | *762W (+ 279aa) |
| SNP | **A** | T | T | Sugar transporter | *133R (+ 133aa) |
| SNP | **C** | T | C | Hypothetical Protein | E168K |
| SNP | **C** | T | T | Protease (S41 family) | T161I |
| SNP | **G** | A | G | B-galactosidase (lacZ_2) | no change |
| DEL | **CT** | C | C | Hypothetical Protein | R65E (+ 9aa) |

**Figure 1.**
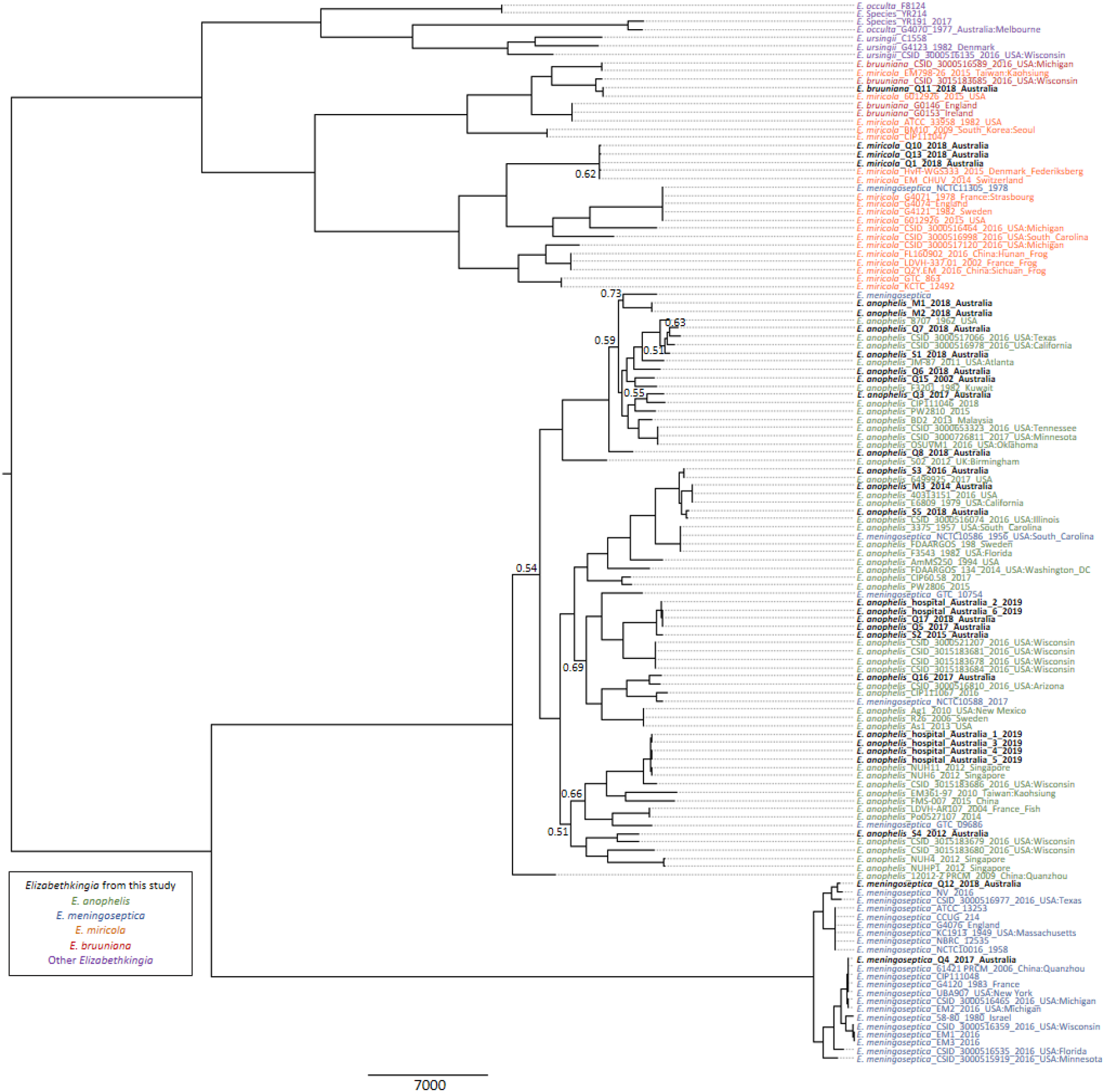
Global phylogenomic analysis of *Elizabethkingia* spp. genomes. Maximum parsimony midpoint-rooted phylogeny. Branches returning bootstrap support <0.8 are labelled. This phylogeny was reconstructed using 127,236 bialleleic, orthologous single-nucleotide polymorphisms identified among the 128 *Elizabethkingia* genomes. Consistency index = 0.4066.

**Figure 2.**
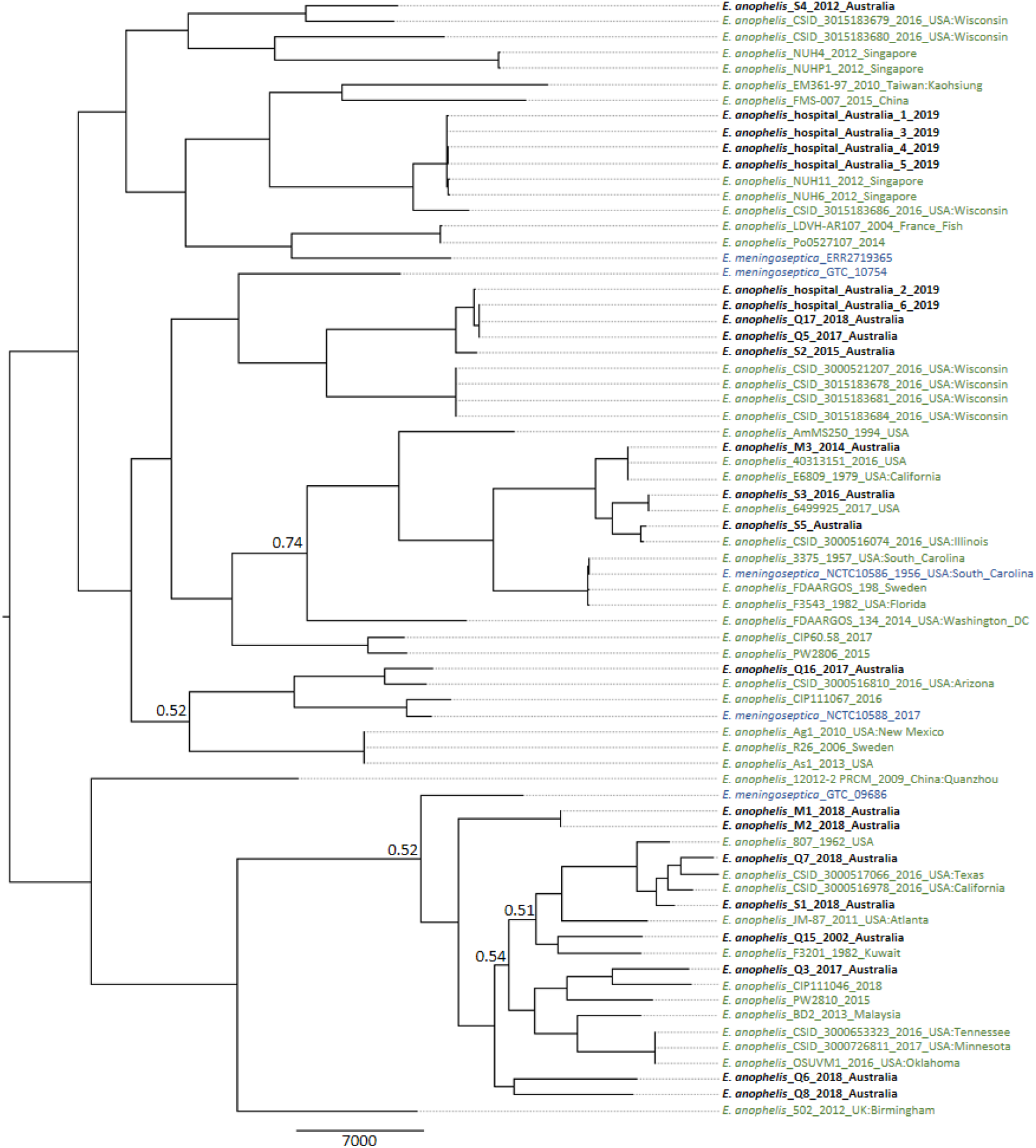
*Elizabethkingia anophelis* species specific phylogenomic analysis. Maximum parsimony midpoint-rooted phylogeny was reconstructed using 121,827 bialleleic, orthologous single-nucleotide polymorphisms identified among the 71 *Elizabethkingia anophelis* genomes. Correctly speciated *Elizabethkingia anophelis* genomes are coloured green, incorrectly speciated *Elizabethkingia meningoseptica* genomes are coloured blue and new *Elizabethkingia anophelis* genomes generated in this study are coloured black. Bootstrap support <0.8 are labelled. Consistency index = 0.3110.

**Figure 3.**
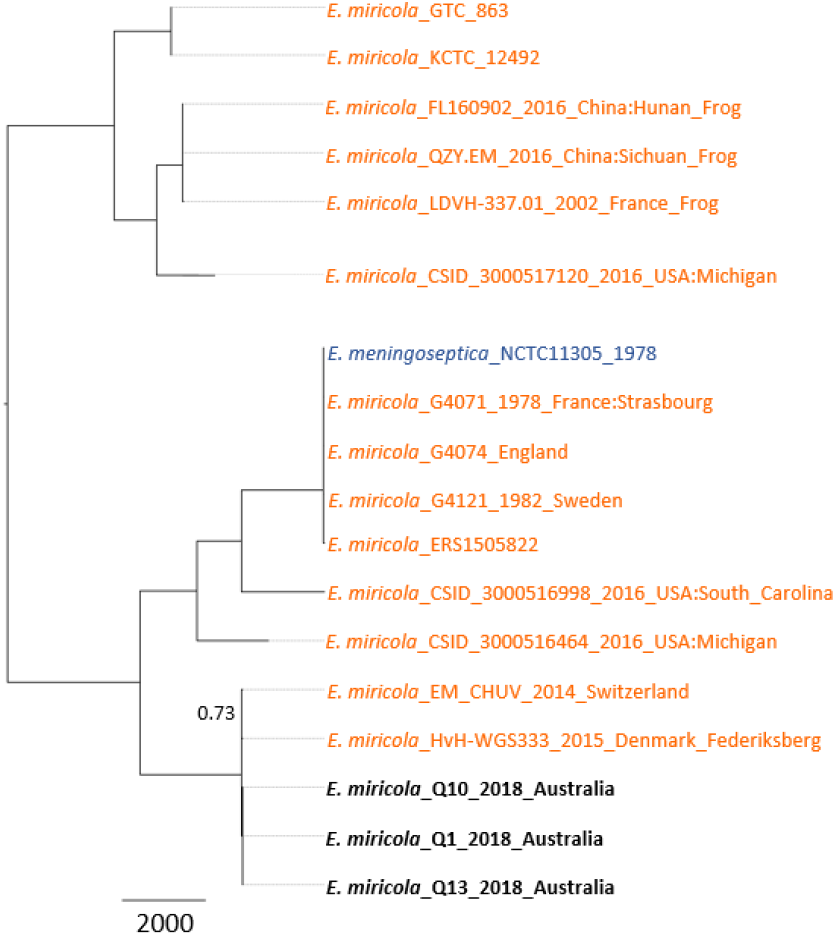
*Elizabethkingia miricola* species specific phylogenomic analysis. Maximum parsimony midpoint-rooted phylogeny was reconstructed using 135,087 bialleleic, orthologous single-nucleotide polymorphisms identified among the 18 genomes. Correctly speciated *E. miricola* strains are coloured orange, incorrectly speciated *Elizabethkingia meningoseptica* coloured blue and *Elizabethkingia anophelis* genomes generated in this study coloured black. Bootstrap support <0.80 is shown. Consistency index = 0.7404.

**Figure 4.**
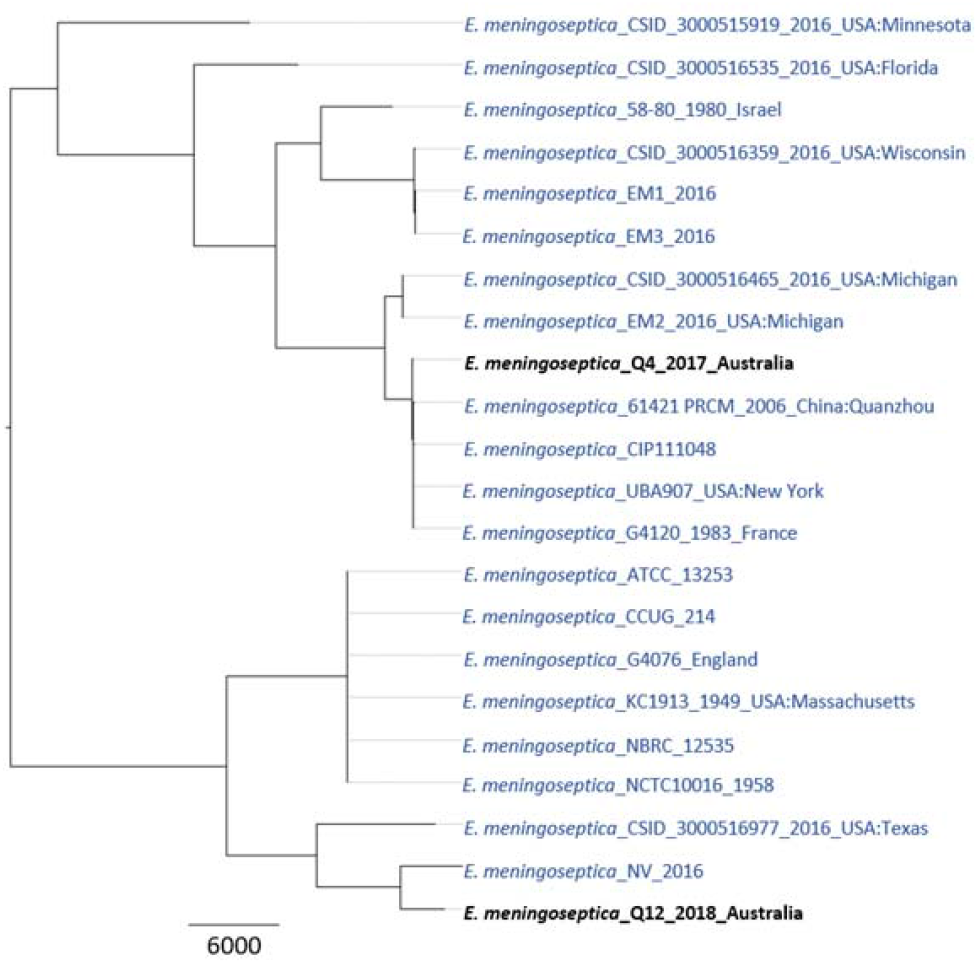
*Elizabethkingia meningoseptica* species specific phylogenomic analysis. Maximum parsimony midpoint-rooted phylogeny was reconstructed using 61,500 bialleleic, orthologous single-nucleotide polymorphisms identified among the 22 genomes. Reference *Elizabethkingia meningoseptica* strains are coloured blue with strains generated in this study coloured black. Bootstrap support is 100 for all branches. Consistency index = 0.6895.

**Figure 5.**
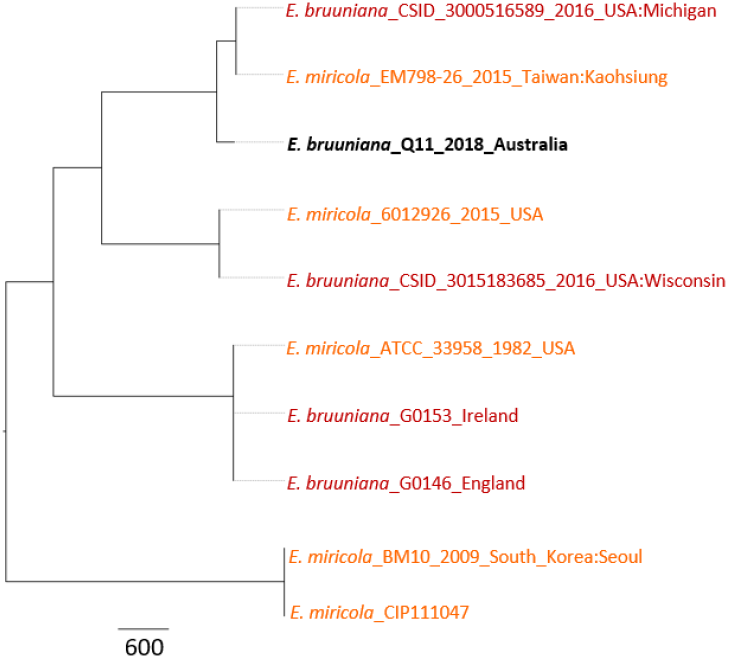
*Elizabethkingia bruuniana* species specific phylogenomic analysis. Maximum parsimony midpoint-rooted phylogeny was reconstructed using 82,680 bialleleic, orthologous single-nucleotide polymorphisms identified among the 10 genomes. Correctly speciated *Elizabethkingia bruuniana* strains are coloured red, incorrectly speciated *Elizabethkingia miricola* strains are coloured orange and genomes generated in this study are coloured black. Bootstrap support <0.8 is shown. Consistency index = 0.8729.

### Minimum Inhibitory Concentration (MIC) testing

*Elizabethkingia* spp. clinical isolates were subjected to broth microdilution to determine MICs for 39 clinically relevant antimicrobials consistent with or complementary to previous *Elizabethkingia* studies^12,23,28,42^ (Tables 3 & 4). Custom Gram-negative Sensititre MIC Plates (ThermoFisher Scientific, Scoresby, VIC, Australia) were used according to manufacturer’s instructions. *E. bruuniana* isolate, EkQ11, was excluded from MIC analyses due to poor growth. *Elizabethkingia* spp. isolates were compared against the European Committee on Antimicrobial Susceptibility Testing (EUCAST) pharmacokinetic-pharmacodynamic (PK-PD) “non-species” breakpoints^43^ and the non-*Enterobacteriaceae* breakpoints as per the Clinical and Laboratory Standards Institute (CLSI) M45 guidelines^44–46^.

**Table 3.**
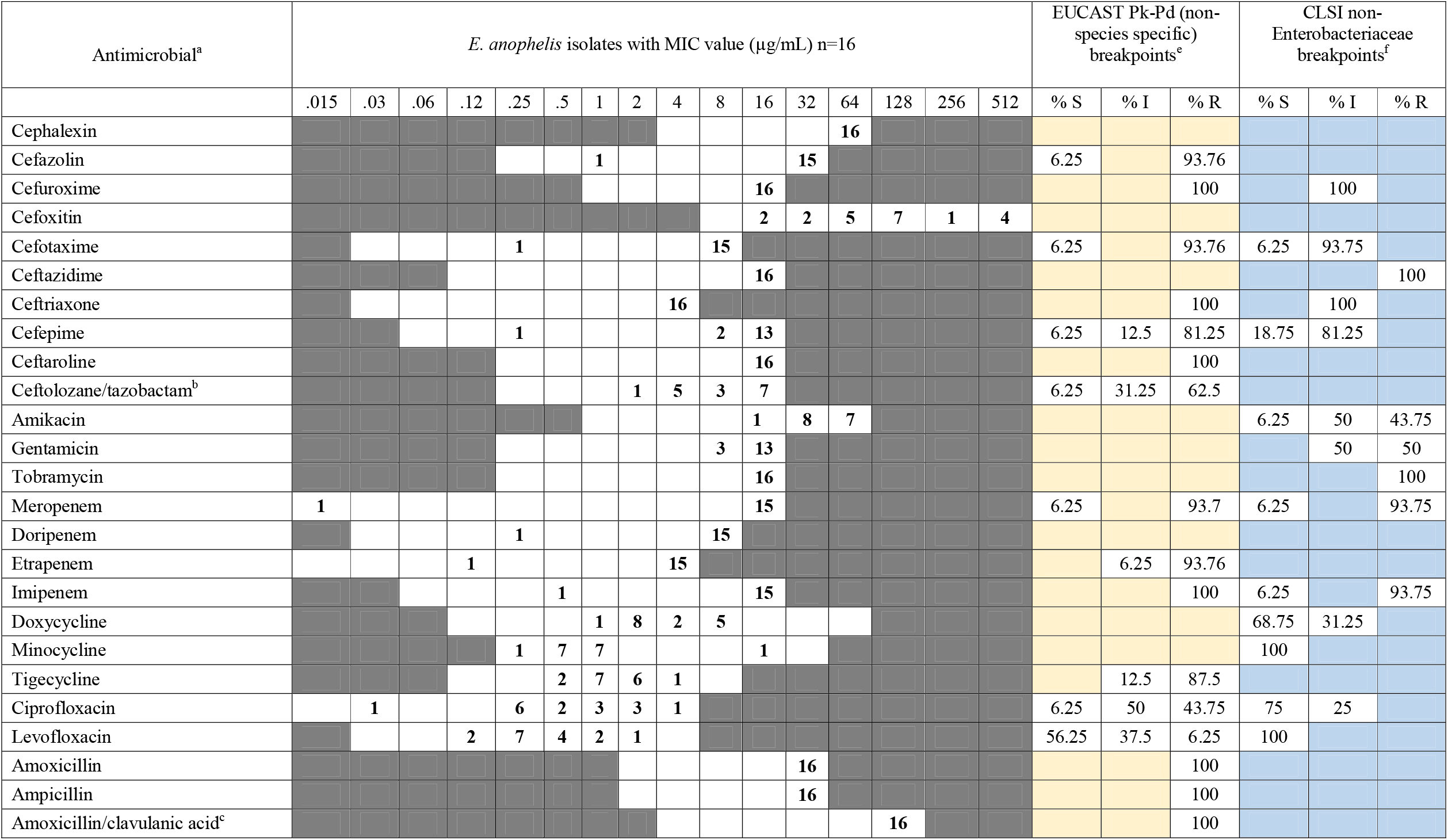
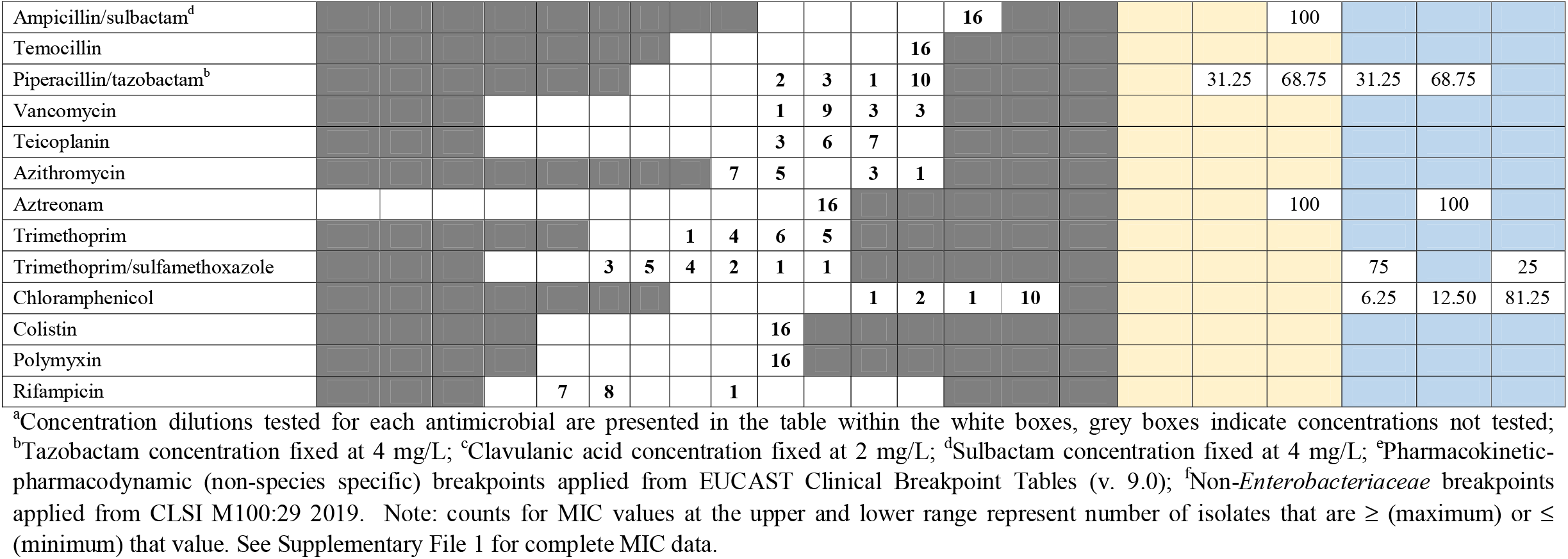
Minimum inhibitory concentration data derived from broth microdilution testing of the 16 Australian *Elizabethkingia anophelis* clinical isolates against 39 clinically relevant antimicrobials. White cells represent the range of concentration tested for each antimicrobial. EUCAST and CLSI breakpoints are shown on the right where available. Blue and yellow cells indicate no breakpoint is currently available for this antimicrobial.

**Table 4.**
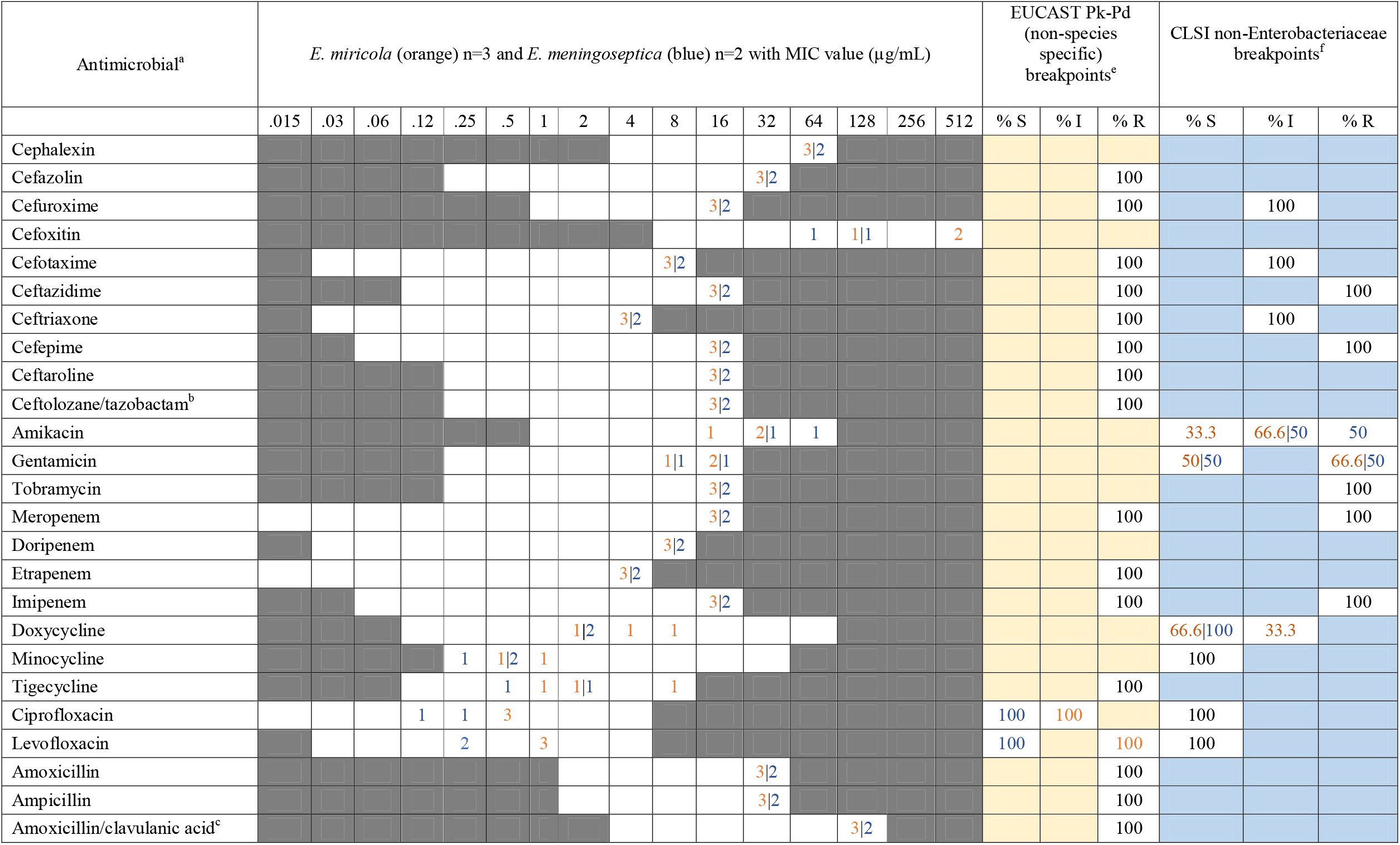
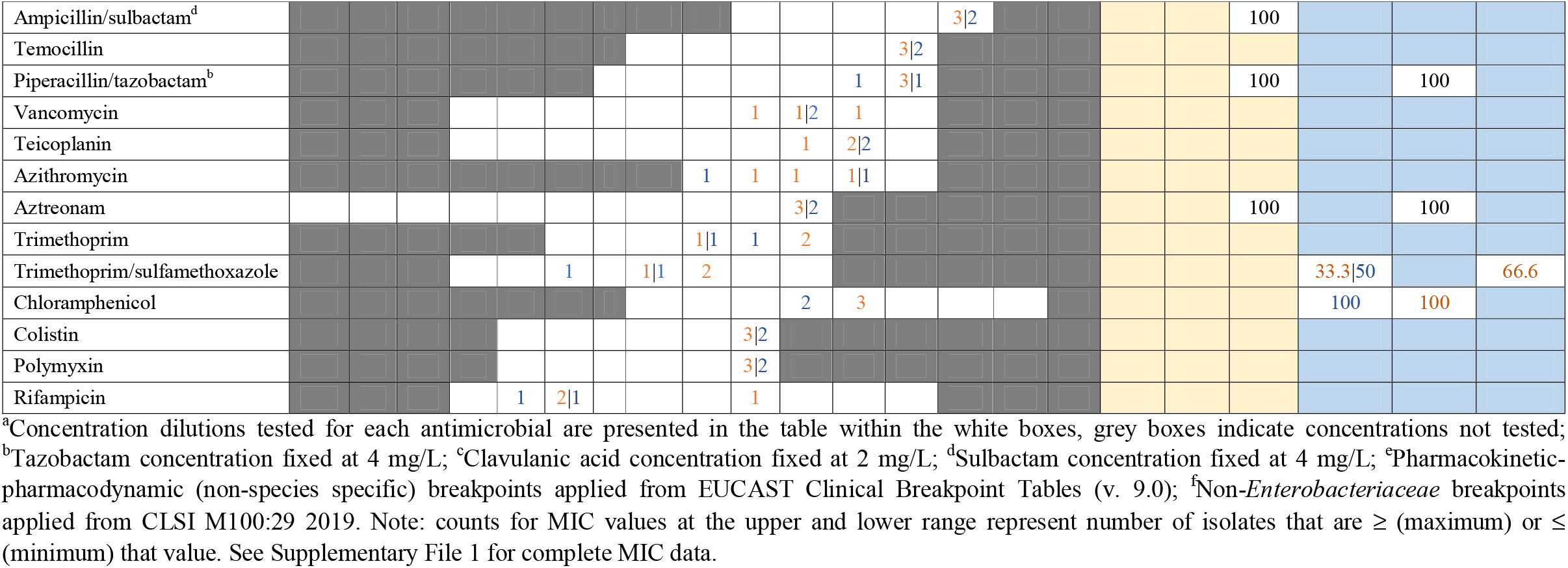
Minimum inhibitory concentration data derived from broth microdilution testing of the 2 *Elizabethkingia meningoseptica* (blue) and 3 *Elizabethkingia miricola* (orange) Australian clinical isolates against 39 clinically relevant antimicrobials. White cells represent the range of concentration tested for each antimicrobial. EUCAST and CLSI breakpoints are shown on the right where available. Blue and yellow cells indicate no breakpoint is currently available for this antimicrobial.

### In silico antimicrobial resistance (AMR) gene predictions

Clinical *Elizabethkingia* spp. WGS data were subject to ABRicate set to the CARD database to predict AMR genes (https://github.com/tseemann/abricate) and RAST for a secondary confirmation^42,47,48^. Geneious prime 2019.2.1 and BLAST (https://blast.ncbi.nlm.nih.gov/Blast.cgi) were used to generate single protein sequence alignments^41^.

### Data availability

Illumina sequence data for the 28 *Elizabethkingia* spp. genomes described in this study have been deposited in the NCBI SRA database under identifier SRP225137, BioProject PRJNA576977 (BioSample accessions: SAMN13016226-SAMN13016247 and SAMN14081590-SAMN14081595).

## Results

### Elizabethkingia speciation using comparative genomics vs mass spectrometry

Phylogenomic reconstruction of 100 *Elizabethkingia* reference genomes collected globally over the past 50 years and the 28 Australian clinical and environmental *Elizabethkingia* spp. genomes robustly identified as *E. anophelis* (*n*=22), *E. miricola* (*n*=3), *E. meningoseptica* (*n*=2) and *E. bruuniana* (*n*=1) (Figure 1; Table S1). Eleven speciation errors were identified in the publicly available dataset consisting of two speciation errors within the *E. anophelis* clade, five within the *E. bruuniana* clade and one within the *E. miricola* clade (Figure 1). Additionally, comparison of the VITEK^®^ MS Knowledge Base v3.2 with genomic species assignments of the Australian isolates resulted in one speciation error in this study, incorrectly identifying *E. bruuniana* as *E. miricola* (Table 1).

### Australian Elizabethkingia and global relatedness

Australian *Elizabethkingia* spp. displayed no distinct phylogeographical signal within the genus phylogeny as they disseminated across the phylogenetic tree (Figure 1.) However, multiple introduction events appear to have taken place, as at least five clades with Australian representatives are branching with international strains, for example: EkQ1, 10 &13 branching with HvH-WGS333 and EM_CHUV from Denmark and Switzerland respectively, EkS4 branching with CSID_3015183679 from Wisconsin, environmental strains EK1,3,4,5 branching with NUH11 and 6 from Singapore, EkQ15 branching with F3201 from Kuwait and EkQ4 clustering with 61421PRCM, G4120 and UBA907 from China, France and New York, respectively (Figure 1). No Australian *Elizabethkingia* isolate was identical to a previously described isolate, with those appearing to be near identical in the phylogenies separated by 16-284 SNPs (Figures 1-5). Australian *E. anophelis* are not closely related to Wisconsin, USA outbreak strains (Figure 1, Figure 2). Clinical isolates EkQ17, Q5, S2 and environmental isolates EK2 and EK6 branched off the Wisconsin, USA outbreak cluster, diverging as a distantly related unique lineage separated by an estimated 20,400 SNPs and 500 indels using CSID_3015183681 as the reference strain. The truncation of the C-terminal of MutY and MutS, characteristic of the outbreak and hypermutator strains were not evident in Australian strains in the amino acid alignment. The 2019 hospital environmental isolates EK1, EK3, EK4, and EK5, collected from various wards handwashing sinks or toilet environments from the same hospital as EkQ5-EkQ17-EK6-EK2, are closely related to two 2012 Singaporean isolates, NUH6 and NUH11. These isolates are separated by 656-867 SNPs and 41-72 indels, and all share a clade with 2016 outbreak isolate CSID_3015183686, which differs from the Singapore isolates by an estimated 9800 SNPs and 260 indels.

### Evidence of E. anophelis nosocomial transmission

Two instances of recent closely related Australian *E. anophelis* isolates were identified on two separate lineages by phylogenetic analysis (Figure 2), both with bootstrap support of 1. In the first instance EkM1 and EkM2 were collected from the same patient one month apart, branching as unique lineage with clinical isolate EkQ6 from a patient in a different hospital. All strains were collected in 2018 and did not show evidence of within host evolution (Figure 2).

In the second instance, diverging from the Wisconsin outbreak cluster in the *E. anophelis* phylogeny are five epidemiologically linked clinical isolates EkQ5, EkQ17, EkS2 and environmental isolates EK2 and EK6 (Figure 2). SNP and indel comparisons between clinical strains EkQ5 and EkQ17 revealed a difference of eight SNPs and one indel between two different patients admitted into the same transplant ward nine months apart in 2018. Epidemiologically, these three isolates appear to be linked to a single environmental source within the transplant ward.

Mutational differences between EkQ5-EkQ17 and EK6 were mostly non-synonymous in nature, consistent with adaptive evolution. Of the two SNPs separating EkQ17 and EK6, one resulted in a missense (E168K) mutation in a hypothetical protein (Ek00046). Between EkQ5 and EK6, four SNPs resulted in missense mutations, and two caused nonsense mutations in the penicillin-binding protein E (PbpE) and a sugar transporter protein that increased protein length, likely leading to altered or lost protein function (Table 2). In addition, the indel mutation accrued by EkQ5 resulted in a frameshift mutation that elongated hypothetical protein (Ek02802) by nine residues, potentially altering its function.

Another hospital environmental isolate, EK2, was linked to the EkQ5-EkQ17-EK6 clade according to phylogenetic analysis, differing by 38 SNPs and 16 indels (Figure 2). This isolate was collected in 2019 from a sink drain in the infectious disease ward adjacent to the transplant ward where EkQ5, EkQ17 and EK6 were isolated. A more distantly related clinical isolate, EkS2, also clustered within the same clade as the EkQ5-EkQ17-EK6-EK2 isolates but differed from these isolates by 3552 SNPs and 120 indels. Consistent with the phylogenomic findings, EkS2 was not epidemiologically linked to the EkQ5-EkQ17-EK6-EK2 isolates, being isolated from a patient admitted to a different hospital in 2015.

### Minimum Inhibitory Concentrations (MIC)

A total of 39 clinically relevant antimicrobials were tested across the 22 clinical *E. anophelis, miricola* and *meningoseptica* isolates. Modal MICs were relatively consistent within and between species and predominantly sat on the higher end of the ranges tested (Tables 3 & 4). *Elizabethkingia* does not have a defined clinical breakpoint, therefore species were examined against the EUCAST “non-species” and CLSI “non-Enterobacteriaceae” PK-PD breakpoints. EUCAST breakpoints suggest Australian strains have the greatest resistance to cephalosporins, carbapenems and penicillins even in combination with β-lactamase inhibitors (amoxicillin-clavulanic acid, piperacillin-tazobactam and ampicillin-sulbactam). Furthermore, the CLSI breakpoints suggest high levels of resistance to amikacin, gentamicin, tobramycin and chloramphenicol. From the MIC values (Tables 3 & 4), only a select few antimicrobials had modal MICs in the lower range, including tetracyclines (doxycycline 2 µg/mL and minocycline 0.5-1 µg/mL), fluoroquinolones (ciprofloxacin 0.25 µg/mL and levofloxacin 0.25 µg/mL) and trimethoprim-sulfamethoxazole 1 µg/mL (Tables 3 & 4). Only minocycline achieved 100% susceptibility across all *E. anophelis* strains using the CLSI non-Enterobacteriaceae PK-PD breakpoints. Rifampicin and azithromycin do not have corresponding EUCAST or CLSI PK-PD breakpoints; however, their respective modal MICs are also on the lower end of the ranges tested, suggesting the potential for susceptibility. One *E. anophelis* isolate EkQ6 was responsible for the low MICs observed across the antimicrobials tested, remaining susceptible to cephalosporins and carbapenems, in addition to the fluoroquinolones, tetracyclines and trimethoprim-sulfamethoxazole.

### In silico antimicrobial resistance (AMR) genes

All 22 clinical *Elizabethkingia* spp. genomes carried all three previously described β-lactamases characteristic of *Elizabethkingia*. The chromosomal extended spectrum β-lactamase *bla*_CME_ encodes cephalosporin and β-lactamase activity, while metallo-β-lactamases *bla*_BlaB_, and *bla*_GOB_ encode activity against carbapenems and β-lactam/β-lactamase inhibitor combinations. The metallo-β-lactamase *bla*_BlaB_, carried a missense mutation of *bla*_BlaB_^ΔT16A^ in EkQ6. Except for *E. bruuniana* EkQ11, all Australian *Elizabethkingia* spp. genomes carried the vancomycin resistance protein VanW. Three *E. miricola* and the *E. bruuniana* isolate carried an AmpC variant with 94-95% sequence similarity to AmpC identified in *E. anophelis* and *E. miricola* genomes (accession numbers CP006576, CP007547 and CP011059). All isolates carried a conserved AmpG, with three strains exhibiting AmpG^ΔM1-A243^ and one strain exhibiting AmpG^ΔM1-A3^ 5’ truncations.

## Discussion

*Elizabethkingia* spp. have caused serious nosocomial infections and outbreaks globally yet have received little attention to date. This study aimed to fill knowledge gaps surrounding diversity, origin and transmission events of clinical and environmental *Elizabethkingia* spp. isolates from Australia, a previously unstudied geographic, using comparative genomics. In parallel, we also describe the antimicrobial resistance profiles among Australian clinical *Elizabethkingia* spp. isolates from broth microdilution data against 39 antimicrobials, to further increase our understanding of suitable treatment options.

### Elizabethkingia speciation using comparative genomics vs mass spectrometry

The 28 Australian *Elizabethkingia* isolates were identified as *E. anophelis, E. meningoseptica, E. miricola* and *E. bruuniana*, with *E. anophelis* as the primary infecting species in Australia, similar to recent global reports^7,22,25^. These isolates, from a previously under-represented geographic area, contribute to ∼20% of the diversity seen in the current reference genome database. Despite previous review of identification failing using mass spectroscopy for species other than *E. anophelis* and *E. meningoseptica*^4^, the VITEK^®^ MS Knowledge Base v3.2 performed reliably in this study with 96.2% accuracy. *E. bruuniana* was the only *Elizabethkingia* species that could not be accurately identified, instead identified as the sister species *E. miricola*. This could be due to the species not yet being present in the database, or perhaps *E. miricola* and *E. bruuniana* being variations of the same species, as many previous speciation errors were seen in the genus phylogeny (Figure 1). Nevertheless, identification of *E. miricola* should be taken with caution until the database has been upgraded with the capabilities to differentiate between the sister-species.

### Australian Elizabethkingia and global relatedness

Our Australian clinical isolates were unique, yet still closely related in comparison to the geographically dispersed reference *Elizabethkingia* spp. genomes (Figures 2-5). The Australian isolates were well dispersed throughout their respective species-specific phylogenies branching with geographically diverse isolates from both clinical and environmental settings. Recently, DNA–DNA hybridization and average nucleotide identity have allowed for the re-classification of *E. miricola* strains ATCC 33958, BM10, and EM798-26 to *E. bruuniana*^3,25,49^. Further to these corrections, using comparative genomics we suggest the re-classification of *E. miricola* strains 6012926 and CIP111047 to *E. bruuniana, E. meningoseptica* strains NCTC10588 and NCTC10586 to *E. anophelis* and lastly, *E. meningoseptica* NCTC11305 to *E. miricola* (Figure 1). Evidence from past studies have described the structure of *E. anophelis* phylogenies to comprise of two and six lineages^6,50^, in this study we identified six lineages, yet as sampling continues this may expand (Figure 2).

Several *E. anophelis* isolates from this study cluster with the Wisconsin outbreak strains from 2016, the most pathogenic *Elizabethkingia* outbreak to date^6^. Outbreak and hypermutator stains have been characterised by their ICE insertion and truncations at the C terminal in both the MutS and MutY protein sequences respectively^6^. The MutS and MutY protein sequences in our clinical isolates aligned with few non-synonymous amino acid changes and no truncations, therefore it is unlikely the Australian clinical isolates would display the outbreak or hypermutator phenotype, which could be responsible for the increased pathogenesis of the Wisconsin strains. Pathogenicity islands were identified in both Australian and Wisconsin *E. anophelis* strains, suggesting they may play an important role in the species survival or pathogenesis.

Hospital environmental isolates EK1, 3, 4, 5 formed a clonal cluster and were closely related to two 2012 Singaporean clinical isolates, NUH6 and NUH11. The Australian environmental isolates differed from the Singaporean isolates by 656-867 SNPs and 41-72 indels suggesting shared ancestry.

### Potential nosocomial transmission of E. anophelis in a transplant ward

A recent case of hospital acquired *E. anophelis* infection was suggested by the identification of a clonal cluster comprised of clinical and environmental isolates in this study. A pair of Australian *E. anophelis* clinical isolates EkQ5 and EkQ17, collected almost a year apart in 2018 from two patients on the transplant ward were characterised as differing by only eight SNPs and one deletion. Additionally, it was found that the hospital environmental sample collected from a hand washing sink in the same transplant ward in late 2019 only differed to clinical sample EkQ5 by six of the above SNPs and the one deletion. The combination of clinical and environmental genomic data, with such low genetic diversity suggests these strains were transmitted via the common reservoir of the hand-washing sink given the extended time frame between patient infection and environmental collection. Near identical isolates have been described previously within *E. anophelis*, such as environmentally collected OSUVM-1 and 2 isolates^51^, hospital outbreak strains NUHP^52^ and Wisconsin CSID^6^ strains, suggesting low genetic variation is not unusual amongst *E. anophelis* infections. The relatedness of sink or toilet environment hospital isolates EK4 and EK5 from the transplant ward, to EK1 and EK3 in the oncology ward suggest that another transmission event may have also taken place, despite not identifying a related clinical isolate. Previous studies have reported contaminated communal water sources as a reservoir for *Elizabethkingia* spp. infections within hospitals^1,17^, with hand-washing stations in a paediatric intensive care unit the source of several *Elizabethkingia* spp. infections in Singapore, where staff transmitted the infection after handwashing^2^. Although, direct human-to-human transmission is seen in many other nosocomial infections^53,54^ and vertical transmission has been reported in *E. anophelis*^55^ the role human-to-human transmission has in *Elizabethkingia* infections still remains unclear. However, given the severity of infection, known patient risk factors and the suggested longevity of the bacteria in the environment, the potential for horizontal transmission should not be overlooked.

### Minimum Inhibitory Concentrations (MIC) testing

The MIC data generated in this study confirm the Australian clinical *Elizabethkingia* spp. isolates (with the exception of isolate EkQ6), like those in previous studies, are resistant to many antimicrobial classes, including cephalosporins, carbapenems and aminoglycosides (Tables 3 & 4)^12,23,24,28,56,56^. From the literature, there is very little variation in *E. anophelis* antimicrobial resistance profiles among isolates from America, Southeast Asia and South Korea. For example, approximately 75-100% of *E. anophelis* isolates were reported as resistant to trimethoprim-sulfamethoxazole^6,23–25,28^, while 75% of Australian strains remained susceptible. Additionally, 88-95% of isolates were susceptible to piperacillin-tazobactam^6,23,25,28^, while 68-70% of Australian and South Korean^24^ isolates were resistant. Vancomycin has been suggested as potential therapy for *E. meningoseptica* infections, therefore we screened our *E. anophelis* strains against vancomycin and additional antimicrobials with Gram-positive activity, such as teicoplanin. Despite some advocating for vancomycin use in *Elizabethkingia* infections^4,29–31^, our data shows resistance within Australian clinical isolates, as MIC values were on the high end of the range tested and all isolates, with the exception of *E. bruuniana* (EkQ11), carry the *vanW* gene. This is the first set of MIC data for teicoplanin and, with a modal MIC of 32 µg/mL, these strains appear to be resistant. Similar to that of the Wisconsin outbreak strains^6^, Australian *Elizabethkingia* spp. strains may be susceptible to azithromycin, as the modal MIC of 4 µg/mL is on the lower end of the range tested. Although doxycycline is not often tested on *E. anophelis* in the literature, unlike in our study, others have found their strains highly susceptible^28^. EUCAST breakpoints suggest 6.25% and 43.75% of Australian *E. anophelis* isolates are resistant to levofloxacin and ciprofloxacin respectively. Variability in fluoroquinolone susceptibility has also been observed in the majority of southeast Asian and American strains^6,23,25,28,31^. Numerous antimicrobials have been tested across *E. anophelis* isolates in previous studies, although susceptibility to multiple antimicrobial classes like that observed in EkQ6, has not been reported previously. Further testing of *E. anophelis* isolates from Australia and abroad would determine if this type of sensitivity is unique to a subset of Australian strains or is present globally.

### In Silico Antimicrobial Resistance (AMR) genes

Antimicrobial resistance genes *bla*_*BlaB*_, *bla*_*GOB*_ and *bla*_*CME*_ were identified within the genomes of all clinical *Elizabethkingia* spp., linking directly to their observed MIC profiles. All isolates with the exception of EkQ6 were resistant to cephalosporins and penicillins (*bla*_CME_), carbapenems and β-lactam/β-lactamase inhibitor combinations (*bla*_BlaB_, and *bla*_GOB_)^4,57–59^. However, fluoroquinolone resistance varied in our collection, as described above. Previous studies have described resistance being mediated by a single step amino acid substitution (Ser83Ile or Ser83Arg) in *gyrA*^23,25,60^, which was not identified in any of the clinical *Elizabethkingia* sp. isolates. The absence of the mutation has also been reported recently for a single isolate in Taiwan^28^. Previous studies have linked DNA topoisomerase IV to an assistance type role in fluoroquinolone resistance for *Elizabethkingia* spp.^28,60^, although this was not identified in our clinical isolate collection either.

In addition, clinical *E. anophelis* isolate EkQ6 carried several mutations not commonly described in proteins BlaB and TopA^25,28,30,61^, yet remained susceptible to cephalosporins, carbapenems, tetracyclines and fluoroquinolones. The substitutions and deletions respectively, may or may not be linked to the susceptibility of this isolate. The observed susceptibility in EkQ6 could have occurred from in-host adaption, evolution in an environment where exposure to antimicrobials is minimal or mutations that have inadvertently resulted in an adaption to a susceptible phenotype. Comparative genomics including more susceptible isolates such as EkQ6 would provide great insight into the intrinsic antimicrobial resistance mechanisms of *Elizabethkingia* species^30,62,63^.

### Potential antimicrobial therapy for Elizabethkingia spp

As *Elizabethkingia* spp. are predominantly isolated from the bloodstream and possess chromosomally encoded MBL-type carbapenemases, therapy is guided by multiple factors such as patient condition prior to infection, the severity and source of infection, previous exposure to antimicrobials and individualised MIC data. In this study, Australian isolates appear to be susceptible to fluoroquinolones, tetracyclines and trimethoprim-sulfamethoxazole. Only levofloxacin and minocycline demonstrated 100% susceptibility using CLSI PK-PD breakpoints. Fluoroquinolone treatment alone has proven to be successful in *Elizabethkingia* spp. infections^64^, yet some recommend combination therapy^65^ in order to mitigate high-level fluoroquinolone resistance for those susceptible to single step mutations. From our and other studies, susceptibility is clearly strain dependent. Our findings suggests rifampicin^66^ or azithromycin could also be effective antimicrobials, although this would require further testing. With this in mind and the recent success of newer antimicrobials against MDR Gram-negative bacteria^67–69^, it would be of value to further test *Elizabethkingia* spp. against newer antimicrobials such as cefiderocol^70^. Although sporadic, *Elizabethkingia* spp. infections have the potential for high mortality rates and nosocomial outbreaks with few treatment options, therefore additional antimicrobial therapies are required and should be investigated further.

## Conclusions

This study has characterised the diversity of Australian *Elizabethkingia* spp. using comparative genomics and antimicrobial resistance genotypically and phenotypically. We have revealed significant strain diversity within Australia and have shown that the VITEK^®^ MS Knowledge Base v3.2 can accurately identify *E. anophelis, E. meningoseptica* and *E. miricola* species, but is yet to correctly identify *E. bruuniana*. Furthermore, genomic exploration has provided insight into the breadth of the intrinsic MDR nature of *Elizabethkingia* spp. and revealed a potential reservoir for infection within a hospital setting where two patients were infected with near identical strains. Antimicrobial resistance data suggests that clinical isolates are susceptible to fluoroquinolones, tetracyclines and trimethoprim-sulfamethoxazole. In particular, minocycline and levofloxacin showed suitable efficacy against *Elizabethkingia* isolates *in vitro*, although further clinical studies are required to define optimal therapy.

## Data Availability

Illumina sequence data for the 28 Elizabethkingia spp. genomes described in this study have been deposited in the NCBI SRA database under identifier SRP225137, BioProject PRJNA576977 (BioSample accessions: SAMN13016226-SAMN13016247 and SAMN14081590- SAMN14081595).

https://www.ncbi.nlm.nih.gov/bioproject/PRJNA576977

## Acknowledgements

https://www.ncbi.nlm.nih.gov/bioproject/PRJNA576977The authors wish to acknowledge the Study Education and Research Committee of Pathology Queensland (LG), University of the Sunshine Coast (DB), Advance Queensland (AQRF13016-17RD2 for DSS; AQIRF0362018 for EPP), and the National Health and Medical Research Council (GNT1157530 for PNAH) for funding this study. We would like to express our gratitude to Mater Pathology, Sullivan and Nicolaides Pathology, and Pathology Queensland for their involvement and support in this project. Finally, we would like to thank the infection control nurses at participating hospitals for environmental sampling.

## Supplementary Material

Table S1. Metadata and ID of the 100 *Elizabethkingia* NCBI and SRA reference genomes used in this study.

Table S2: SPAdes and prokka genome assembly and annotation statistics of Australian

*Elizabethkingia* clinical isolates analysed in this study.

## References

1. Moore, L. S. P. et al. Waterborne Elizabethkingia meningoseptica in Adult Critical Care1. Emerg. Infect. Dis. 22, 9–17 (2016).

2. Yung, C.-F. et al. Elizabethkingia anophelis and Association with Tap Water and Handwashing, Singapore. Emerg. Infect. Dis. 24, 1730–1733 (2018).

3. Nicholson, A. C. et al. Revisiting the taxonomy of the genus Elizabethkingia using whole-genome sequencing, optical mapping, and MALDI-TOF, along with proposal of three novel Elizabethkingia species: Elizabethkingia bruuniana sp. nov., Elizabethkingia ursingii sp. nov., and Elizabethkingia occulta sp. nov. Antonie Van Leeuwenhoek 111, 55–72 (2018).

4. Lin, J.-N., Lai, C.-H., Yang, C.-H. & Huang, Y.-H. Elizabethkingia Infections in Humans: From Genomics to Clinics. Microorganisms 7, 295 (2019).

5. Nicholson, A. C., Humrighouse, B. W., Graziano, J. C., Emery, B. & McQuiston, J. R. Draft Genome Sequences of Strains Representing Each of the Elizabethkingia Genomospecies Previously Determined by DNA-DNA Hybridization. Genome Announc. 2.

6. Perrin, A. et al. Evolutionary dynamics and genomic features of the Elizabethkingia anophelis 2015 to 2016 Wisconsin outbreak strain. Nat. Commun. 8, 15483 (2017).

7. Chew, K. L., Cheng, B., Lin, R. T. P. & Teo, J. W. P. Elizabethkingia anophelis Is the Dominant Elizabethkingia Species Found in Blood Cultures in Singapore. J. Clin. Microbiol. 56, (2018).

8. Jean, S. S., Lee, W. S., Chen, F. L., Ou, T. Y. & Hsueh, P. R. Elizabethkingia meningoseptica: an important emerging pathogen causing healthcare-associated infections. J. Hosp. Infect. 86, 244–249 (2014).

9. Frank, T. et al. First case of Elizabethkingia anophelis meningitis in the Central African Republic. Lancet Lond. Engl. 381, 1876 (2013).

10. Issack, M. I. & Neetoo, Y. An outbreak of Elizabethkingia meningoseptica neonatal meningitis in Mauritius. J. Infect. Dev. Ctries. 5, 834–839 (2011).

11. Teo, J. et al. First case of E. anophelis outbreak in an intensive-care unit. The Lancet 382, 855–856 (2013).

12. Hsu, M.-S. et al. Clinical features, antimicrobial susceptibilities, and outcomes of Elizabethkingia meningoseptica (Chryseobacterium meningosepticum) bacteremia at a medical center in Taiwan, 1999–2006. Eur. J. Clin. Microbiol. Infect. Dis. 30, 1271–1278 (2011).

13. Choi, M. H. et al. Risk Factors for Elizabethkingia Acquisition and Clinical Characteristics of Patients, South Korea. Emerg. Infect. Dis. 25, 42–51 (2019).

14. Janda, J. M. & Lopez, D. L. Mini review: New pathogen profiles: Elizabethkingia anophelis. Diagn. Microbiol. Infect. Dis. 88, 201–205 (2017).

15. Weaver, K. N. et al. Acute emergence of Elizabethkingia meningoseptica infection among mechanically ventilated patients in a long-term acute care facility. Infect. Control Hosp. Epidemiol. 31, 54–58 (2010).

16. Chawla, K., Gopinathan, A., Varma, M. & Mukhopadhyay, C. Elizabethkingia Meningoseptica Outbreak in Intensive Care Unit. J. Glob. Infect. Dis. 7, 43–44 (2015).

17. Kyritsi, M. A., Mouchtouri, V. A., Pournaras, S. & Hadjichristodoulou, C. First reported isolation of an emerging opportunistic pathogen (Elizabethkingia anophelis) from hospital water systems in Greece. J. Water Health 16, 164–170 (2018).

18. Doijad, S., Ghosh, H., Glaeser, S., Kämpfer, P. & Chakraborty, T. Taxonomic reassessment of the genus Elizabethkingia using whole-genome sequencing: Elizabethkingia endophytica Kämpfer et al. 2015 is a later subjective synonym of Elizabethkingia anophelis Kämpfer et al. 2011. Int. J. Syst. Evol. Microbiol. 66, 4555–4559 (2016).

19. Li, Y. et al. Chryseobacterium miricola sp. nov., A Novel Species Isolated from Condensation Water of Space Station Mir. Syst. Appl. Microbiol. 26, 523–528 (2003).

20. Kampfer, P. et al. Elizabethkingia anophelis sp. nov., isolated from the midgut of the mosquito Anopheles gambiae. Int. J. Syst. Evol. Microbiol. 61, 2670–2675 (2011).

21. Jorgensen, J. H. & P. Faller, M. A. Introduction to the 11th Edition of the Manual of Clinical Microbiology. Man. Clin. Microbiol. Elev. Ed. 3–4 (2015) doi:10.1128/9781555817381.ch1.

22. Lau, S. K. P. et al. Elizabethkingia anophelis bacteremia is associated with clinically significant infections and high mortality. Sci. Rep. 6, (2016).

23. Lin, J.-N., Lai, C.-H., Yang, C.-H., Huang, Y.-H. & Lin, H.-H. Clinical manifestations, molecular characteristics, antimicrobial susceptibility patterns and contributions of target gene mutation to fluoroquinolone resistance in Elizabethkingia anophelis. J. Antimicrob. Chemother. 73, 2497–2502 (2018).

24. Han, M.-S. et al. Relative Prevalence and Antimicrobial Susceptibility of Clinical Isolates of Elizabethkingia Species Based on 16S rRNA Gene Sequencing. J. Clin. Microbiol. 55, 274–280 (2017).

25. Lin, J.-N., Lai, C.-H., Yang, C.-H. & Huang, Y.-H. Comparison of Clinical Manifestations, Antimicrobial Susceptibility Patterns, and Mutations of Fluoroquinolone Target Genes between Elizabethkingia meningoseptica and Elizabethkingia anophelis Isolated in Taiwan. J. Clin. Med. 7, (2018).

26. Lin, Y.-T. et al. Clinical and microbiological analysis of Elizabethkingia meningoseptica bacteremia in adult patients in Taiwan. Scand. J. Infect. Dis. 41, 628–634 (2009).

27. Tai, I.-C. et al. Outbreak of Elizabethkingia meningoseptica sepsis with meningitis in a well-baby nursery. J. Hosp. Infect. 96, 168–171 (2017).

28. Jian, M.-J. et al. Fluoroquinolone resistance in carbapenem-resistant Elizabethkingia anophelis: phenotypic and genotypic characteristics of clinical isolates with topoisomerase mutations and comparative genomic analysis. 8 (2019).

29. Jean, S.-S., Hsieh, T.-C., Ning, Y.-Z. & Hsueh, P.-R. Role of vancomycin in the treatment of bacteraemia and meningitis caused by Elizabethkingia meningoseptica. Int. J. Antimicrob. Agents 50, 507–511 (2017).

30. Chang, T.-Y., Chen, H.-Y., Chou, Y.-C., Cheng, Y.-H. & Sun, J.-R. In vitro activities of imipenem, vancomycin, and rifampicin against clinical Elizabethkingia species producing BlaB and GOB metallo-beta-lactamases. Eur. J. Clin. Microbiol. Infect. Dis. (2019) doi:10.1007/s10096-019-03639-3.

31. Han, M.-S. et al. Relative Prevalence and Antimicrobial Susceptibility of Clinical Isolates of Elizabethkingia Species Based on 16S rRNA Gene Sequencing. J. Clin. Microbiol. 55, 274–280 (2017).

32. Bolger, A. M., Lohse, M., & Usadel, B. Trimmomatic: A flexible trimmer for Illumina Sequence Data. Bioinforma. Btu 170 (2014).

33. Ewels, P., Magnusson, M., Lundin, S. & Käller, M. MultiQC: summarize analysis results for multiple tools and samples in a single report. Bioinformatics 32, 3047–3048 (2016).

34. H. Li. Seqtk: a fast and lightweight tool for processing FASTA or FASTQ sequences. (2013).

35. Huang, W., Li, L., Myers, J. R. & Marth, G. T. ART: a next-generation sequencing read simulator. Bioinformatics 28, 593–594 (2012).

36. Nurk, S. et al. Assembling Genomes and Mini-metagenomes from Highly Chimeric Reads. in Research in Computational Molecular Biology (eds. Deng, M., Jiang, R., Sun, F. & Zhang, X.) 158–170 (Springer Berlin Heidelberg, 2013).

37. Seemann, T. Prokka: rapid prokaryotic genome annotation. Bioinforma. Oxf. Engl. 30, 2068–2069 (2014).

38. Sarovich, D. S. & Price, E. P. SPANDx: a genomics pipeline for comparative analysis of large haploid whole genome re-sequencing datasets. BMC Res. Notes 7, 618 (2014).

39. Swofford, D. L. PAUP* ver 4.0. b10. Phylogenetic Anal. Using Parsimony Methods Sunderland MA Sinauer Assoc. Sunderland (2003).

40. Using Tablet for visual exploration of second-generation sequencing data | Briefings in Bioinformatics | Oxford Academic. https://academic.oup.com/bib/article/14/2/193/208254.

41. Kearse, M. et al. Geneious Basic: an integrated and extendable desktop software platform for the organization and analysis of sequence data. Bioinforma. Oxf. Engl. 28, 1647–1649 (2012).

42. Aziz, R. K. et al. The RAST Server: rapid annotations using subsystems technology. BMC Genomics 9, 75 (2008).

43. Kahlmeter, G. et al. European Committee on Antimicrobial Susceptibility Testing (EUCAST) Technical Notes on antimicrobial susceptibility testing. Clin. Microbiol. Infect. 12, 501–503 (2006).

44. Clinical and Laboratory Standards Institute. CLSI guideline M45. Methods for Antimicrobial Dilution and Disk Susceptibility Testing of Infrequently Isolated or Fastidious Bacteria. 3rd ed, Wayne, PA: Clinical and Laboratory Standards Institute; 2016.

45. Jorgensen, J. H., Hindler, J. F., Reller, L. B. & Weinstein, M. P. New Consensus Guidelines from the Clinical and Laboratory Standards Institute for Antimicrobial Susceptibility Testing of Infrequently Isolated or Fastidious Bacteria. Clin. Infect. Dis. 44, 280–286 (2007).

46. Clinical and Laboratory Standards Institute. 2015. M7-A10: methods for dilution antimicrobial susceptibility tests for bacteria that grow aerobically; approved standard, 10th ed. Clinical and Laboratory Standards Institute, Wayne, PA.

47. Brettin, T. et al. RASTtk: a modular and extensible implementation of the RAST algorithm for building custom annotation pipelines and annotating batches of genomes. Sci. Rep. 5, 8365 (2015).

48. Overbeek, R. et al. The SEED and the Rapid Annotation of microbial genomes using Subsystems Technology (RAST). Nucleic Acids Res. 42, D206–214 (2014).

49. Lin, J.-N., Lai, C.-H., Yang, C.-H., Huang, Y.-H. & Lin, H.-H. Genomic features, phylogenetic relationships, and comparative genomics of Elizabethkingia anophelis strain EM361-97 isolated in Taiwan. Sci. Rep. 7, (2017).

50. Breurec, S. et al. Genomic epidemiology and global diversity of the emerging bacterial pathogen Elizabethkingia anophelis. Sci. Rep. 6, (2016).

51. Johnson, W. L. et al. The draft genomes of Elizabethkingia anophelis of equine origin are genetically similar to three isolates from human clinical specimens. PLOS ONE 13, e0200731 (2018).

52. Teo, J. et al. Comparative Genomic Analysis of Malaria Mosquito Vector-Associated Novel Pathogen Elizabethkingia anophelis. Genome Biol. Evol. 6, 1158–1165 (2014).

53. Tagoe, D. N. A., Baidoo, S., Dadzie, I., Tengey, D. & Agede, C. Potential Sources of Transmission of Hospital Acquired Infections in the Volta Regional Hospital in Ghana. Ghana Med. J. 45, 22–26 (2011).

54. Khan, H. A., Baig, F. K. & Mehboob, R. Nosocomial infections: Epidemiology, prevention, control and surveillance. Asian Pac. J. Trop. Biomed. 7, 478–482 (2017).

55. Lau, S. K. P. et al. Evidence for Elizabethkingia anophelis transmission from mother to infant, Hong Kong. Emerg. Infect. Dis. 21, 232–241 (2015).

56. Lin, J.-N., Lai, C.-H., Yang, C.-H., Huang, Y.-H. & Lin, H.-H. Genomic Features, Comparative Genomics, and Antimicrobial Susceptibility Patterns of Elizabethkingia bruuniana. Sci. Rep. 9, 2267 (2019).

57. Bellais, S., Poirel, L., Naas, T., Girlich, D. & Nordmann, P. Genetic-biochemical analysis and distribution of the Ambler class A beta-lactamase CME-2, responsible for extendedspectrum cephalosporin resistance in Chryseobacterium (Flavobacterium) meningosepticum. Antimicrob. Agents Chemother. 44, 1–9 (2000).

58. Rossolini, G. M. et al. Cloning of a Chryseobacterium (Flavobacterium) meningosepticum chromosomal gene (blaA(CME)) encoding an extended-spectrum class A beta-lactamase related to the Bacteroides cephalosporinases and the VEB-1 and PER beta-lactamases. Antimicrob. Agents Chemother. 43, 2193–2199 (1999).

59. Colapietro, M. et al. BlaB-15, a new BlaB metallo-beta-lactamase variant found in an Elizabethkingia miricola clinical isolate. Diagn. Microbiol. Infect. Dis. 85, 195–197 (2016).

60. Jian, M.-J., Cheng, Y.-H., Perng, C.-L. & Shang, H.-S. Molecular typing and profiling of topoisomerase mutations causing resistance to ciprofloxacin and levofloxacin in Elizabethkingia species. PeerJ 6, (2018).

61. Wang, M. et al. The antibiotic resistance and pathogenicity of a multidrug-resistant Elizabethkingia anophelis isolate. MicrobiologyOpen 0, e804.

62. Opota, O. et al. Genome of the carbapenemase-producing clinical isolate Elizabethkingia miricola EM_CHUV and comparative genomics with Elizabethkingia meningoseptica and Elizabethkingia anophelis: evidence for intrinsic multidrug resistance trait of emerging pathogens. Int. J. Antimicrob. Agents 49, 93–97 (2017).

63. Balm, M. N. et al. Bad design, bad practices, bad bugs: frustrations in controlling an outbreak of Elizabethkingia meningoseptica in intensive care units. J Hosp Infect 85, 134–40 (2013).

64. Huang, Y.-C., Lin, Y.-T. & Wang, F.-D. Comparison of the therapeutic efficacy of fluoroquinolone and non-fluoroquinolone treatment in patients with Elizabethkingia meningoseptica bacteraemia. Int. J. Antimicrob. Agents 51, 47–51 (2018).

65. Chan, J. C. et al. Invasive paediatric Elizabethkingia meningoseptica infections are best treated with a combination of piperacillin/tazobactam and trimethoprim/sulfamethoxazole or fluoroquinolone. J. Med. Microbiol. 68, 1167–1172 (2019).

66. Bassetti, M., Peghin, M., Vena, A. & Giacobbe, D. R. Treatment of Infections Due to MDR Gram-Negative Bacteria. Front. Med. 6, (2019).

67. Hackel, M. A. et al. In Vitro Activity of the Siderophore Cephalosporin, Cefiderocol, against Carbapenem-Nonsusceptible and Multidrug-Resistant Isolates of Gram-Negative Bacilli Collected Worldwide in 2014 to 2016. Antimicrob. Agents Chemother. 62, (2018).

68. Haidar, G. et al. Ceftolozane-Tazobactam for the Treatment of Multidrug-Resistant Pseudomonas aeruginosa Infections: Clinical Effectiveness and Evolution of Resistance. Clin. Infect. Dis. Off. Publ. Infect. Dis. Soc. Am. 65, 110–120 (2017).

69. Shields, R. K. et al. Verification of Ceftazidime-Avibactam and Ceftolozane-Tazobactam Susceptibility Testing Methods against Carbapenem-Resistant Enterobacteriaceae and Pseudomonas aeruginosa. J. Clin. Microbiol. 56, (2018).

70. Zhanel, G. G. et al. Cefiderocol: A Siderophore Cephalosporin with Activity Against Carbapenem-Resistant and Multidrug-Resistant Gram-Negative Bacilli. Drugs 79, 271–289 (2019).

